# Unmet need for family planning and associated factors among currently married women in Nepal: A further analysis of Nepal Demographic and Health Survey - 2022

**DOI:** 10.1101/2023.08.16.23294154

**Authors:** Saugat Pratap KC, Bikram Adhikari, Achyut Raj Pandey, Merina Pandey, Sampurna Kakchapati, Santosh Giri, Shreeman Sharma, Bipul Lamichhane, Ghanshyam Gautam, Deepak Joshi, Bishnu Prasad Dulal, Shophika Regmi, Sushil Chandra Baral

## Abstract

**Introduction:** Family planning (FP) is crucial for improving maternal and newborn health outcomes, promoting gender equality, and reducing poverty. Unmet FP needs persist globally, especially in South Asia and Sub-Saharan Africa leading to unintended pregnancies, unsafe abortions, and maternal fatalities. Nepal, despite efforts, faces significant unmet FP needs, particularly among specific groups. This study aims to identify the determinants of unmet needs for FP from a nationally representative survey.

**Methods:** We analyzed the data from nationally representative Nepal Health Demographic Survey 2022. There was a total of 14,845 participants in the survey selected through systematic random sampling technique. Weighted analysis using R statistical software was conducted binary and multinomial logistic regression were performed to assess associations between independent variables and unmet FP needs.

**Results:** The total unmet FP need was 20.8% (95%CI: 19.7, 21.9) accounting 13.4% (95%CI: 12.5, 14.4) for unmet need for limiting and 7.4% (95%CI: 6.8, 8.0) for unmet for spacing. In multivariable regression model, age, ethnicity, religion, participants and partners education level, province, occupation, wealth quintile, parity and desire for child were statistically associated with total unmet need of FP. Lower odds of total unmet need for FP were present in 20-34 years and 35-49 years compared to <20 years, women belonging to Madhesi ethnic group(AOR: 0.79, 95% CI:0.65, 0.97) compared to brahmin/Chhetri, women from richest (AOR: 0.73, 95% CI:0.59, 0.90) and middle wealth quintile (AOR: 0.83, 95% CI:0.70, 0.99) group compared poorest wealth quintile and women belonging to rural area (AOR: 0.89, 95%CI: 0.80, 0.99) compared to urban area. Whereas higher odds of total unmet need for FP were present in non-Hindu women (AOR: 1.30, 95% CI:1.10, 1.53) compared to Hindu women and women belonging to Madhesh, Lumbini and Sudurpaschim province compared to Bagmati province.

**Conclusion:** Nepal faces relatively high unmet FP needs across various socio-demographic strata. Addressing these needs requires targeted interventions focusing on age, ethnicity, religion, education, and socio-economic factors to ensure universal access to FP services.

## Introduction

Family planning (FP) is a crucial aspect of reproductive health, enabling individuals to make informed choices about when to have children and how many to have^1^. Ensuring universal access to FP is a fundamental human right that plays a central role in promoting gender equality and empowering women. Additionally, it serves as a crucial factor in alleviating poverty and advancing the objective of achieving Universal Health Coverage (UHC)^2^. The World Health Organization (WHO) defines unmet FP needs as fecund and sexually active women who desire to postpone or limit childbearing but are not using any contraceptive method^3^. In 2019, despite the various advantages of FP and efforts to enhance accessibility, approximately 160 million women and adolescents worldwide still lacked access to adequate family planning services. More than half of these women with unmet needs resided in Sub-Saharan Africa and South Asia^4^.

The presence of a significant unmet need for FP results in elevated rates of unintended pregnancies, which, in turn, are closely linked to unsafe abortions and maternal fatalities. These connections are well-established in the realm of public health research^5,6^. Addressing the persistent demand for FP has remained a core focus of global health and population strategies for many years^7^. Empowering women to make their own decisions regarding pregnancy, rather than being forced into it, can lead to broader social and economic advantages extending beyond the health sector. These benefits include higher levels of education, increased participation of women in the workforce, and greater accumulation of wealth within households^8^. Addressing the unmet need for FP aligns with two Sustainable Development Goals (SDGs). The SDG 3 focuses on promoting well-being and ensuring universal access to sexual and reproductive health-care services, including the integration of reproductive health into national strategies and programs. The SDG 5 aims to achieve gender equality and empower women and girls by guaranteeing universal access to sexual and reproductive health services and reproductive rights^9^.

Multiple factors have been identified in the literature as crucial elements associated with unmet need for FP among women in developing countries^10^. In Nepal, multiple studies based on the Demographic and Health Survey 2011 have revealed a high prevalence of unmet need for family planning^11,12^, with disparities based on education and age^11^. Additionally, research conducted in Nepal has shown that the proportion of contraceptive users decreases with longer travel times to access family planning outlets, impacting both urban and rural women^13,14^. Women’s societal position and decision-making power are closely intertwined with family planning access and utilization. Despite efforts by leading health organizations to address inequities in unmet need for family planning, the problem persists^15^, indicating the need for further action. Thus, this study aims to explore the unmet need for family planning among reproductive age women in Nepal using the most recent demographic and health survey.

## Methods

### Study design

We analyzed Nepal Health Demographic Survey, 2022 (NDHS 2022) dataset in this study. NDHS 2022 is the nationally representative survey implemented by New ERA under the aegis of the Ministry of Health and Population (MoHP) with the technical support of ICF International. NDHS 2022 was funded by the US Agency for International Development (USAID).

### Study setting

Nepal, positioned in Southeast Asia, is a landlocked country spanning an area of 147,516 km2. It is divided into seven administrative provinces, encompassing a total of 753 municipalities, including 6 metropolitan cities, 11 sub-metropolitan cities, 276 urban municipalities, and 460 rural municipalities. The geographical landscape of Nepal consists of three ecological regions: Mountain, Hill, and Terai. According to the Census of 2021, Nepal’s total population reached 29,164,578 individuals, with females accounting for 14,911,027 (51.1%). The Human Development Index of rural and urban Nepal were 0.647 and 0.561 respectively with overall human development index of 0.587.

### Sample and sampling

The sampling and sampling technique of NDHS 2022 is described elsewhere^16^. The samples of NDHS are nationally representative, encompassing all seven provinces of the nation. The initial stratification involved dividing each province into urban and rural areas, creating a sampling stratum for each province. This led to the establishment of 14 sampling strata in total. The sampling process comprised two stages. Initially, 476 primary sampling units (PSUs) were chosen using a probability-proportional-to-size approach. Out of these, 248 PSUs were from urban regions, and 228 were from rural regions. Subsequently, 30 households were selected from each PSU in the second stage, resulting in an overall sample size of 14,280 households, with 7,440 from urban areas and 6,840 from rural areas. Among the households surveyed, there were 15,283 eligible women aged 15-49 for individual interviews. Interviews were successfully conducted with 14,845 of these women, yielding a response rate of 97%. For the purposes of this study, data from 11,180 currently married women were included.

### Data collection

Data collection for NDHS 2022 occurred between January 5 and June 22, 2022, utilizing 19 teams. Each team was comprised of a supervisor, a male interviewer, three female interviewers, and a biomarker specialist.

### Dependent variables

#### Unmet need of FP

The adolescents women who were not pregnant and not postpartum amenorrheic and are considered fecund and want to postpone their next birth for 2 or more years or stop childbearing altogether but are not using a contraceptive method, or have a mistimed or unwanted current pregnancy, or are postpartum amenorrheic and their most recent birth in the last 2 years was mistimed or unwanted were considered to have unmet need of family planning^21^.

### Independent variables

The independent variables assessed in this study included ecological belt (Mountain/Hill/Terai), setting (Urban/Rural), province(Koshi/Madhesh/Bagmati/Gandaki/Lumbini/Karnali/Sudurpaschim), age (in years), ethnicity (Brahmin or Chhetri/ Dalit/Janajatis/Madhesi/Other), religion (Hindu/Non-Hindu), marital status (Unmarried/Married or living together/Divorced or non-living), wealth quintile (Poorest/Poorer/Middle/Richer/Richest), education(No education/Basic/Secondary/Higher), occupation (Not working/Agriculture/Professional or technical or manager or clerical), and health insurance(Covered/Not covered), media exposure (Present/Not present).

### Statistical analysis

We performed pre-analytical and statistical analysis using R version 4.2.0 and RStudio. We performed a weighted analysis was used using “survey package” to account complex survey design of NDHS 2022. We presented categorical variables as frequency, percent and 95% CI whereas numerical variables as mean and 95% CI. We used univariate and multivariable, binary, and multinomial logistic regression to determine the association of unmet need of FP with independent variables. The results of the regression analysis were presented as crude odds ratio (COR), adjusted odds ratio (AOR), and 95% CI.

### Ethical approval

We requested the DHS program for permission to use NDHS 2022 dataset which was granted to download and use NDHS 2022 dataset from https://www.dhsprogram.com. NHDS 2022 obtained ethical approval from the institutional review board of ICF International, United States of America (Reference number: 180657.0.001.NP.DHS.01, Date: 28^th^ April 2022) and the ethical review board of Nepal Health Research Council (Reference number: 678, Date: 30^th^ September 2021)

## Results

Table 1 show the socio-economic characteristics of married women. More than two-third of women (67.6%) of women resided in urban areas. More than half of the women (56.1%) were from the terai region. Madhesh province had the highest representation at 21.6%, followed by Bagmati (19.3%), Lumbini (18.3%) and Koshi province (16.9%). Slightly more than one-third (36.2%) of the women belonged to Janajati followed by Brahmin/Chettri (27.1%), Madheshi (16.4%), and Dalit (27.1%). Majority (83.5%) of the women were Hindu. The median age of the women is 32.0 years and most of the women (53.7%) belonged to 20-34 years age group. The distribution of married women across wealth quintiles revealed that a significant portion, 21.3% belonged to the richer quintile, followed closely by the middle (20.8%), richest (19.9%), poorer (19.8%), and poorest quintiles (18.2%). About 38% of the women’s husbands had secondary education, while 13.3% were uneducated. According to women’s occupation, more than half (53%) of the surveyed women were engaged in agriculture and 23.9% of women were not working. 49.8% of the women had exposure to media. The majority of women (72.5%) did not desire more children. Most women (66.5%) were multipara, primipara and nullipara women constituted 24.1% and 9.4% respectively.

**Table 1:** Characteristics of Currently married women.

| Characteristic | n | % (95%CI) |
| --- | --- | --- |
| **Place of residence** |  |  |
| Urban | 7,553 | 67.6 (66.1, 69.0) |
| Rural | 3,627 | 32.4 (31.0, 33.9) |
| **Ecological region** |  |  |
| Mountain | 629 | 5.6 (3.92, 8.01) |
| Hill | 4,275 | 38.2 (34.5, 42.1) |
| Terai | 6,276 | 56.1 (52.4, 59.8) |
| **Province** |  |  |
| Koshi | 1,887 | 16.9 (15.7, 18.2) |
| Madhesh | 2,419 | 21.6 (20.4, 23.0) |
| Bagmati | 2,156 | 19.3 (17.7, 21.0) |
| Gandaki | 1,046 | 9.4 (8.32, 10.5) |
| Lumbini | 2,020 | 18.1 (16.9, 19.3) |
| Karnali | 691 | 6.2 (5.66, 6.75) |
| Sudurpashchim | 960 | 8.6 (7.93, 9.30) |
| **Ethnicity** |  |  |
| Brahmin/Chhetri | 3,031 | 27.1 (24.9, 29.5) |
| Dalit | 1,734 | 15.5 (13.7, 17.5) |
| Janajati | 4,042 | 36.2 (33.4, 39.0) |
| Madheshi | 1,835 | 16.4 (14.3, 18.7) |
| Others | 539 | 4.8 (3.42, 6.74) |
| **Religion** |  |  |
| Hindu | 9,338 | 83.5 (81.4, 85.4) |
| Non-Hindu | 1,842 | 16.5 (14.6, 18.6) |
| **Current age (in years)** | 32.0 (25.0, 39.0) | 32.0, 33.0 |
| <20 years | 563 | 5 (4.52, 5.60) |
| 20-34 years | 6,007 | 53.7 (52.6, 54.9) |
| 35-49 years | 4,609 | 41.2 (40.1, 42.4) |
| **Wealth index** |  |  |
| Poorest | 2,031 | 18.2 (16.3, 20.2) |
| Poorer | 2,217 | 19.8 (18.1, 21.7) |
| Middle | 2,323 | 20.8 (19.3, 22.4) |
| Richer | 2,381 | 21.3 (19.7, 23.0) |
| Richest | 2,228 | 19.9 (17.6, 22.5) |
| **Highest educational level** |  |  |
| No education | 3,475 | 31.1 (29.4, 32.9) |
| Basic | 3,701 | 33.1 (31.8, 34.5) |
| Secondary | 3,536 | 31.6 (30.1, 33.2) |
| Higher | 468 | 4.2 (3.59, 4.88) |
| **Husband/partner's education level** |  |  |
| No education | 1,482 | 13.3 (12.1, 14.5) |
| Basic | 4,470 | 40 (38.3, 41.6) |
| Secondary | 4,250 | 38 (36.5, 39.6) |
| Higher | 822 | 7.3 (6.45, 8.36) |
| don't know | 156 | 1.4 (1.11, 1.75) |
| **Occupation** |  |  |
| Not working | 2,677 | 23.9 (22.1, 25.9) |
| Agriculture | 5,920 | 53 (50.1, 55.8) |
| Administrative Clerk | 734 | 6.6 (5.83, 7.38) |
| Sales and service | 928 | 8.3 (7.43, 9.26) |
| Skilled/unskilled labor | 907 | 8.1 (7.24, 9.08) |
| Other | 14 | 0.1 (0.05, 0.28) |
| **Media exposure** | 5,568 | 49.8 (47.8, 51.8) |
| **Desire for child** |  |  |
| Having another | 2,681 | 24 (22.9, 25.1) |
| Undecided | 394 | 3.5 (3.07, 4.05) |
| Wants no more | 8,104 | 72.5 (71.3, 73.7) |
| **Parity** |  |  |
| Multipara | 7,429 | 66.5 (65.3, 67.6) |
| Primipara | 2,696 | 24.1 (23.1, 25.1) |
| Nullipara | 1,055 | 9.4 (8.7, 10.2) |

n: frequency; CI: confidence interval

Table 2 presents unmet need of FP among different subcategories. The unmet need for spacing births is higher in urban areas (7.5%) than in rural areas (7.2%), while the unmet need for limiting births is higher in rural areas (13.9%) compared to urban areas (13.2%). The hill region has the highest unmet need for limiting births (16.1%), while the mountain region has the highest unmet need for spacing births (7.9%). The Gandaki province stands out with the highest total unmet need (28.1%), as well as the highest unmet need for limiting births (20.6%). On the other hand, the Bagmati province has the lowest total unmet need (16%) and the lowest unmet need for spacing births (4.3%). The Dalit ethnic group has the highest total unmet need (25.5%), with the majority of it being unmet need for spacing births (11%). Unmet need for FP is highest among women aged below 20 years (30.9%) and decreases with age. The majority of their unmet need is for spacing births (28.4%). The poorest women have the highest total unmet need (24.7%), while the richest women have the lowest total unmet need (16.9%). Women with no education have the lowest unmet need for spacing births (3.2%) and the highest unmet need for limiting births (13.2%). The unmet need for limiting births is highest among agricultural workers (14.7%) and lowest among. those in sales and service occupations (11.1%).

**Table 2:** Unmet need of family planning among different subcategories.

| Characteristic | n | Total unmet need<br>% (95%CI) | Unmet need for<br>spacing, % (95%CI) | Unmet need<br>for limiting, %<br>(95%CI) |
| --- | --- | --- | --- | --- |
| **Place of residence** |  |  |  |  |
| Urban | 7,553 | 20.7 (19.2, 22.1) | 7.5 (6.66, 8.39) | 13.2 (12.0, 14.4) |
| Rural | 3,627 | 21.1 (19.6, 22.7) | 7.2 (6.41, 7.97) | 13.9 (12.7, 15.3) |
| **Ecological region** |  |  |  |  |
| Mountain | 629 | 19.1 (16.1, 22.5) | 7.9 (6.15, 10.2) | 11.2 (8.99, 13.8) |
| Hill | 4,275 | 22.7 (21.2, 24.4) | 6.6 (5.85, 7.44) | 16.1 (14.7, 17.6) |
| Terai | 6,276 | 19.7 (18.1, 21.3) | 7.8 (6.91, 8.89) | 11.8 (10.6, 13.1) |
| **Province** |  |  |  |  |
| Koshi | 1,887 | 17.6 (15.7, 19.6) | 7.9 (6.48, 9.64) | 9.6 (8.29, 11.2) |
| Madhesh | 2,419 | 21.1 (18.4, 24.1) | 9.9 (8.17, 12.0) | 11.2 (9.30, 13.4) |
| Bagmati | 2,156 | 16 (14.0, 18.2) | 4.3 (3.36, 5.38) | 11.7 (9.89, 13.9) |
| Gandaki | 1,046 | 28.1 (24.8, 31.5) | 7.4 (5.94, 9.29) | 20.6 (17.7, 23.9) |
| Lumbini | 2,020 | 23.3 (20.3, 26.6) | 6.8 (5.56, 8.39) | 16.5 (14.0, 19.4) |
| Karnali | 691 | 23.4 (20.8, 26.2) | 8.5 (6.89, 10.4) | 14.9 (12.8, 17.3) |
| Sudurpashchim | 960 | 22.1 (19.1, 25.3) | 7.2 (5.99, 8.62) | 14.9 (12.4, 17.8) |
| **Ethnicity** |  |  |  |  |
| Brahmin/Chhetri | 3,031 | 20.7 (18.9, 22.7) | 6.2 (5.33, 7.16) | 14.6 (13.1, 16.2) |
| Dalit | 1,734 | 25.5 (23.3, 28.0) | 11 (9.45, 12.8) | 14.5 (12.5, 16.8) |
| Janajati | 4,042 | 19.7 (18.1, 21.5) | 6.5 (5.66, 7.54) | 13.2 (11.9, 14.7) |
| Madheshi | 1,835 | 17.6 (14.9, 20.5) | 6.7 (5.37, 8.22) | 10.9 (8.76, 13.5) |
| Others | 539 | 24.8 (19.3, 31.2) | 11.1 (7.22, 16.7) | 13.7 (10.4, 17.8) |
| **Religion** |  |  |  |  |
| Hindu | 9,338 | 20.4 (19.3, 21.6) | 7.1 (6.45, 7.77) | 13.3 (12.4, 14.3) |
| Non-Hindu | 1,842 | 22.8 (20.2, 25.6) | 8.8 (7.05, 11.0) | 13.9 (12.0, 16.1) |
| Current age (in years), Median (IQR) | - | 32.0 (25.0, 39.0) | 33.0 (28.0, 39.0) | 23.0 (20.0, 26.0) |
| <20 years | 563 | 30.9 (26.8, 35.4) | 28.4 (24.3, 32.8) | 2.6 (1.62, 4.02) |
| 20-34 years | 6,007 | 24.7 (23.3, 26.1) | 10.8 (9.89, 11.8) | 13.9 (12.8, 15.0) |
| 35-49 years | 4,609 | 14.5 (13.2, 15.9) | 0.3 (0.17, 0.66) | 14.2 (12.9, 15.5) |
| **Wealth index** |  |  |  |  |
| Poorest | 2,031 | 24.7 (22.7, 26.8) | 8.7 (7.48, 10.0) | 16.1 (14.4, 17.8) |
| Poorer | 2,217 | 21.4 (19.3, 23.6) | 8.6 (7.40, 10.0) | 12.7 (11.1, 14.6) |
| Middle | 2,323 | 20.4 (18.5, 22.6) | 7.3 (6.19, 8.52) | 13.2 (11.6, 15.0) |
| Richer | 2,381 | 20.9 (18.9, 23.1) | 7.7 (6.28, 9.37) | 13.3 (11.6, 15.1) |
| Richest | 2,228 | 16.9 (14.5, 19.6) | 4.7 (3.81, 5.86) | 12.2 (10.0, 14.7) |
| **Highest educational level** |  |  |  |  |
| No education | 3,475 | 16.4 (14.7, 18.2) | 3.2 (2.34, 4.34) | 13.2 (11.7, 14.8) |
| Basic | 3,701 | 23.7 (22.1, 25.4) | 8.1 (7.12, 9.15) | 15.6 (14.3, 17.1) |
| Secondary | 3,536 | 22.7 (21.0, 24.6) | 11 (9.81, 12.4) | 11.7 (10.5, 13.1) |
| Higher | 468 | 16.1 (12.0, 21.2) | 5.3 (3.12, 8.75) | 10.8 (7.55, 15.2) |
| **Husband/partner's education level** |  |  |  |  |
| No education | 1,482 | 15.3 (13.0, 18.0) | 4.4 (3.22, 5.98) | 10.9 (9.02, 13.1) |
| Basic | 4,470 | 22.2 (20.7, 23.8) | 7 (6.10, 8.00) | 15.2 (14.0, 16.5) |
| Secondary | 4,250 | 21.8 (20.3, 23.5) | 8.9 (7.88, 9.94) | 13 (11.7, 14.4) |
| Higher | 822 | 16.6 (13.8, 19.8) | 5.5 (4.04, 7.43) | 11.1 (8.88, 13.8) |
| don't know | 156 | 26.1 (18.3, 35.8) | 16 (10.4, 23.9) | 10.1 (6.12, 16.1) |
| **Occupation** |  |  |  |  |
| Not working | 2,677 | 22.7 (20.5, 25.1) | 11.1 (9.45, 13.0) | 11.6 (9.98, 13.5) |
| Agriculture | 5,920 | 21.3 (19.9, 22.7) | 6.5 (5.86, 7.26) | 14.7 (13.6, 16.0) |
| Administrative Clerk | 734 | 17.7 (14.6, 21.2) | 5.8 (4.15, 8.02) | 11.9 (9.21, 15.2) |
| Sales and service | 928 | 15.5 (12.7, 18.8) | 4.4 (3.13, 6.14) | 11.1 (8.85, 13.8) |
| Skilled/unskilled labor | 907 | 20.3 (17.1, 23.9) | 6.2 (4.60, 8.40) | 14.1 (11.5, 17.1) |
| Other | 14 | 3.9 (0.47, 26.1) | 3.9 (0.47, 26.1) | 0 (<0.01, <0.01) |
| **Media exposure** | 5,568 | 20 (18.7, 21.4) | 7 (6.15, 7.87) | 13.1 (12.0, 14.3) |
| **Desire for child** |  |  |  |  |
| Having another | 2,681 | 21.9 (20.0, 23.9) | 21.9 (20.0, 23.9) | 0 |
| Undecided | 394 | 32.4 (26.9, 38.5) | 32.4 (26.9, 38.5) | 0 |
| Wants no more | 8,104 | 19.9 (18.7, 21.1) | 1.4 (1.11, 1.65) | 18.5 (17.3, 19.8) |
| **Parity** |  |  |  |  |
| Multipara | 7,429 | 19.7 (18.4, 21.1) | 2.8 (2.28, 3.32) | 17 (15.7, 18.3) |
| Primipara | 1,055 | 16 (13.6, 18.8) | 15.6 (13.2, 18.3) | 0.5 (0.17, 1.23) |
| Nullipara | 2,696 | 25.7 (23.8, 27.7) | 16.9 (15.2, 18.7) | 8.8 (7.63, 10.1) |
| **Total** | 11,180 | 20.8 (19.7, 21.9) | 13.4 (12.5, 14.4) | 7.4 (6.8, 8.0) |

*n*: frequency; *CI*: confidence interval

**Table 3:** Factors associated with unmet need for FP.

|  | Total unmet need |  | Unmet needs for limiting |  | Unmet need for spacing |  |
| --- | --- | --- | --- | --- | --- | --- |
|  | COR (95%CI) | AOR (95%CI) | COR (95%CI) | AOR (95%CI) | COR (95%CI) | AOR (95%CI) |
| **Age** |  |  |  |  |  |  |
| <20 years | Ref | Ref | Ref | Ref | Ref | Ref |
| 20-34 years | 0.73 (0.61, 0.88)** | 0.60 (0.48, 0.75)** | 4.97 (2.92, 8.46)** | 1.62 (0.90, 2.93) | 0.35 (0.29, 0.43)** | 0.61 (0.48, 0.78)** |
| 35-49 years | 0.38 (0.31, 0.46)** | 0.35 (0.27, 0.44)** | 4.47 (2.63, 7.62)** | 1.02 (0.56, 1.86) | 0.01 (0.01, 0.02)** | 0.07 (0.04, 0.12)** |
| **Ethnicity** |  |  |  |  |  |  |
| Brahmin/Chhetri | Ref | Ref | Ref | Ref | Ref | Ref |
| Dalit | 1.31 (1.14, 1.51)** | 1.14 (0.97, 1.35) | 1.06 (0.90, 1.26) | 1.09 (0.89, 1.33) | 1.89 (1.53, 2.34)** | 1.31 (0.99, 1.74) |
| Janajati | 0.94 (0.84, 1.06) | 0.89 (0.78, 1.03) | 0.9 (0.78, 1.03) | 0.95 (0.81, 1.12) | 1.04 (0.86, 1.27) | 0.8 (0.63, 1.03) |
| Madheshi | 0.81 (0.70, 0.94) | 0.79 (0.65, 0.97)* | 0.72 (0.60, 0.86)** | 0.93 (0.73, 1.18) | 1.04 (0.82, 1.31) | 0.66 (0.47, 0.93)* |
| Others | 1.26 (1.02, 1.56) | 1.03 (0.76, 1.41) | 0.99 (0.76, 1.29) | 1.31 (0.89, 1.93) | 1.89 (1.39, 2.58)** | 0.82 (0.49, 1.37) |
| **Religion** |  |  |  |  |  |  |
| Hindu | Ref | Ref | Ref | Ref | Ref | Ref |
| Non-Hindu | 1.15 (1.02, 1.30)* | 1.30 (1.10, 1.53)* | 1.08 (0.93, 1.25) | 1.28 (1.05, 1.56)* | 1.29 (1.07, 1.54) | 1.37 (1.03, 1.82)* |
| **Highest educational level** |  |  |  |  |  |  |
| No education | Ref | Ref | Ref | Ref | Ref | Ref |
| Basic | 1.59 (1.41, 1.78)** | 1.35 (1.18, 1.55)** | 1.3 (1.14, 1.48)** | 1.29 (1.11, 1.51)** | 2.77 (2.22, 3.47)** | 1.65 (1.24, 2.21)** |
| Secondary | 1.5 (1.33, 1.69)** | 1.39 (1.18, 1.65)** | 0.96 (0.83, 1.11) | 1.27 (1.04, 1.56)* | 3.73 (3.01, 4.64)** | 1.91 (1.38, 2.64)** |
| Higher | 0.98 (0.75, 1.27) | 1.26 (0.89, 1.76) | 0.82 (0.60, 1.11) | 1.35 (0.90, 2.03) | 1.64 (1.05, 2.57)* | 1.5 (0.82, 2.74) |
| **Occupation** |  |  |  |  |  |  |
| Not Working | Ref | Ref | Ref | Ref | Ref | Ref |
| Agriculture | 0.92 (0.82, 1.02) | 0.97 (0.85, 1.10) | 1.24 (1.08, 1.43) | 1.03 (0.87, 1.21) | 0.58 (0.49, 0.68)** | 0.96 (0.79, 1.18) |
| Profes_tech_manag_cler | 0.73 (0.59, 0.90) | 0.88 (0.70, 1.11) | 0.96 (0.74, 1.23) | 0.95 (0.71, 1.26) | 0.49 (0.35, 0.68)** | 0.88 (0.59, 1.31) |
| Sales and Service | 0.62 (0.51, 0.76)** | 0.7 (0.56, 0.86)** | 0.87 (0.69, 1.11) | 0.73 (0.57, 0.94)* | 0.36 (0.26, 0.51)** | 0.63 (0.43, 0.92)* |
| Skille or unskilled labor | 0.87 (0.72, 1.04) | 0.94 (0.77, 1.14) | 1.17 (0.94, 1.46) | 1.01 (0.80, 1.29) | 0.54 (0.40, 0.73)** | 0.87 (0.62, 1.22) |
| Other | 0.14 (0.01, 2.14) | 0.21 (0.01, 3.28) |  |  | 0.28 (0.02, 4.38) | 2.63 (0.09, 74.42) |
| ***Wealth Index Quintile*** |  |  |  |  |  |  |
| Poorest | Ref | Ref | Ref | Ref | Ref | Ref |
| Poorer | 0.83 (0.72, 0.95) | 0.88 (0.75, 1.03) | 0.76 (0.64, 0.90)* | 0.83 (0.68, 1.01) | 0.95 (0.77, 1.18) | 0.92 (0.70, 1.20) |
| Middle | 0.78 (0.68, 0.90)** | 0.83 (0.70, 0.99)* | 0.78 (0.65, 0.92)* | 0.88 (0.71, 1.08) | 0.79 (0.64, 0.99) | 0.72 (0.54, 0.97)* |
| Richer | 0.81 (0.70, 0.93)* | 0.85 (0.71, 1.02) | 0.79 (0.66, 0.93)* | 0.89 (0.71, 1.11) | 0.84 (0.68, 1.05) | 0.76 (0.56, 1.05) |
| Richest | 0.62 (0.53, 0.72)** | 0.73 (0.59, 0.90)* | 0.69 (0.58, 0.82)** | 0.78 (0.61, 1.01) | 0.49 (0.38, 0.64)** | 0.62 (0.43, 0.90)* |
| ***Parity*** |  |  |  |  |  |  |
| Multipara | Ref | Ref | Ref | Ref | Ref | Ref |
| Nullipara | 0.78 (0.65, 0.93)* | 0.51 (0.41, 0.64)** | 0.03 (0.01, 0.06)** | 0.2 (0.08, 0.51)** | 5.42 (4.36, 6.73)** | 0.66 (0.50, 0.89) |
| Primipara | 1.41 (1.27, 1.56)** | 1.05 (0.92, 1.21) | 0.56 (0.48, 0.65)** | 0.99 (0.84, 1.18) | 6.63 (5.58, 7.88)** | 1.37 (1.09, 1.72) |
| ***Desire for child*** |  |  |  |  |  |  |
| having another | Ref | Ref |  |  | Ref | Ref |
| Undecided | 1.71 (1.36, 2.15)** | 1.76 (1.39, 2.24)** |  |  | 1.71 (1.36, 2.15)** | 1.92 (1.50, 2.46)** |
| Wants no more | 0.88 (0.79, 0.98)* | 1.14 (0.99, 1.32) |  |  | 0.06 (0.05, 0.07)** | 0.13 (0.10, 0.17)** |
| ***Place of residence*** |  |  |  |  |  |  |
| Urban | Ref | Ref | Ref | Ref | Ref | Ref |
| Rural | 1.03 (0.93, 1.13) | 0.89 (0.80, 0.99)* | 1.06 (0.95, 1.20) | 0.89 (0.77, 1.01) | 0.96 (0.82, 1.12) | 0.88 (0.73, 1.06) |
| ***Ecological region*** |  |  |  |  |  |  |
| Mountain | Ref | Ref | Ref | Ref | Ref | Ref |
| Hill | 1.24 (1.01, 1.54) | 1.23 (0.98, 1.54) | 1.5 (1.16, 1.95)* | 1.45 (1.09, 1.92)* | 0.87 (0.64, 1.20) | 0.88 (0.61, 1.28) |
| Terai | 1.04 (0.84, 1.28) | 0.92 (0.71, 1.18) | 1.06 (0.82, 1.38) | 1.02 (0.74, 1.40) | 1 (0.73, 1.35) | 0.77 (0.50, 1.18) |
| ***Province*** |  |  |  |  |  |  |
| Koshi | Ref | Ref | Ref | Ref | Ref | Ref |
| Madhesh province | 1.26 (1.08, 1.46)* | 1.64 (1.36, 1.99)** | 1.21 (0.99, 1.48) | 1.46 (1.14, 1.87)* | 1.31 (1.05, 1.62) | 1.9 (1.39, 2.58)** |
| Bagmati province | 0.89 (0.76, 1.05) | 0.9 (0.74, 1.09) | 1.19 (0.98, 1.46) | 1.07 (0.85, 1.36) | 0.53 (0.40, 0.69) | 0.64 (0.46, 0.90)* |
| Gandaki province | 1.83 (1.53, 2.19)** | 1.74 (1.42, 2.13)** | 2.45 (1.97, 3.04)** | 2.11 (1.65, 2.70)** | 1.08 (0.81, 1.44) | 1.27 (0.89, 1.82) |
| Lumbini province | 1.43 (1.22, 1.67)** | 1.62 (1.37, 1.92)** | 1.84 (1.51, 2.23)** | 1.98 (1.60, 2.44)** | 0.93 (0.73, 1.18) | 1.11 (0.84, 1.47) |
| Karnali province | 1.43 (1.16, 1.77)** | 1.16 (0.91, 1.47) | 1.66 (1.28, 2.16)** | 1.28 (0.95, 1.72) | 1.15 (0.84, 1.58) | 1.04 (0.70, 1.56) |
| Sudurpashchim province | 1.33 (1.10, 1.61)* | 1.39 (1.13, 1.71)* | 1.63 (1.29, 2.07)** | 1.6 (1.23, 2.06)** | 0.96 (0.71, 1.30) | 1.1 (0.78, 1.57) |
| ***Husband/partner's education level*** |  |  |  |  |  |  |
| No education | Ref | Ref | Ref | Ref | Ref | Ref |
| Basic | 1.58 (1.35, 1.85)** | 1.37 (1.15, 1.63)** | 1.52 (1.26, 1.82)** | 1.46 (1.20, 1.79)** | 1.73 (1.32, 2.28)** | 1.03 (0.73, 1.45) |
| Secondary | 1.55 (1.32, 1.81)** | 1.35 (1.12, 1.64)* | 1.29 (1.07, 1.55) | 1.46 (1.16, 1.82)** | 2.18 (1.66, 2.86)** | 1.01 (0.70, 1.44) |
| Higher | 1.1 (0.87, 1.39) | 1.17 (0.88, 1.55) | 1.04 (0.79, 1.36) | 1.37 (0.98, 1.92) | 1.27 (0.86, 1.88) | 0.82 (0.49, 1.36) |
| Don't know | 1.95 (1.33, 2.87)** | 1.62 (1.09, 2.40) | 1.06 (0.61, 1.83) | 1.2 (0.68, 2.14) | 4.18 (2.54, 6.88)** | 1.95 (1.10, 3.46)* |
| ***Media Exposure*** |  |  |  |  |  |  |
| No | Ref | Ref | Ref | Ref | Ref | Ref |
| Yes | 0.91 (0.83, 1.00)* | 0.92 (0.83, 1.01) | 0.93 (0.83, 1.04) | 0.9 (0.79, 1.01) | 0.88 (0.76, 1.01) | 0.95 (0.80, 1.12) |

*COR: Crude Odds ratio; AOR: Adjusted odds ratio; CI: Confidence interval; Ref: reference group*
*\* Significance at 0.05 level of significance*
*\*\* significance at 0.001 level of significance*

In multivariable logistic regression. the women belonging to 20-34 years and 35-49 years age group have lower odds of total unmet need for FP and unmet need spacing compared to <20 years age group. Women belonging to Madheshi ethnicity had 21% (AOR: 0.79;95%CI: 0.65, 0.97) lower odds of total unmet need for FP and 34% (AOR: 0.66; 95%CI: 0.47, 0.93) lower odds of unmet need for spacing compared to Brahmin/Chettri (AOR: 1.31, 95% CI:0.99, 1,74). Women who were non-Hindus had 28% higher odds to have unmet need for limiting (AOR:1.28, 95% CI: 1.05, 1.56) and 37% higher odds of unmet need for spacing (AOR: 1.37, 95% CI: 1.03, 1.82) compared to Hindu women. The odds of unmet need for spacing was 2-fold higher among women who attained secondary education compared with their counterparts (AOR: 1.91; 95% CI: 1.38, 2.64). Women belonging to wealthier households are less likely to have total unmet need for FP, unmet need for spacing and limiting. Women who were undecided regarding having a child had almost 2 folds higher odds of having unmet need for spacing than women desiring to have another child. (AOR: 1.92, 95% CI: 1.50, 2.46). Women from the hill region were 1.45 times likely to have unmet need for limiting. Women residing in Gandaki province had 2 times higher odds of unmet need for limiting compared with their counterparts (AOR: 2.11, 95% CI: 1.65, 2.70). With an increase in husband’s education, the likelihood of unmet need for spacing and unmet need for limiting tends to fall. Likelihood of unmet need for spacing and unmet need for limiting were lower among women who had mass media exposure.

## Discussion

The overall aim of this study was identifying associated factors determining unmet need for FP among currently married women in Nepal. Among South Asian countries, the unmet need of FP in Nepal is higher than Bangladesh (12% in 207-18), India (9.4% in 2019-21) and Pakistan (17.3% in 2017-19)^17^. This variance might plausibly stem from differences in the accessibility of health services, awareness, and attitudes towards FP provisions, coupled with influences from socioeconomic, demographic, and cultural aspects.

The current study demonstrates that women below the age of 20 exhibit higher unmet need for FP in total and unmet need for spacing compared to their counterparts. These findings resonate with other studies in India and Ethiopia^18–20^. This pattern can potentially be attributed to the relative immaturity of women in this age bracket, leading to challenges in decision-making regarding FP, alongside potential difficulties in overcoming the influences from spouses, in-laws, and the broader community. However, the proportion of unmet need for limiting is higher among women aged 20 to 39Relatively higher unmet need for limiting among 20-34 years seem reasonable, as they may have already achieved their desired/planned number of children and have higher need which often is not met. Conversely, a heightened desire for birth spacing might be projected among younger women, who want to postpone their next pregnancy. Our findings substantiate results from other studies^21–26^.

The findings of our study reveal a notable disparity in unmet need for FP, with rural women having higher unmet need for FP compared to their urban counterparts. These findings resonate with a study conducted in Ethiopia^27^, underscoring the existence of discrepancies in family planning unmet needs between urban and rural contexts. Possible explanation for urban/rural differences could be attributed to cultural and behavioral influences along with far off location of facilities that are not equipped with all methods of FP and services interrupted by lack of commodities and problems in the supply chain could bring about this disparity.

Women belonging to the Dalit Ethnicity have slightly higher odds of unmet need for spacing compared to those from the Brahmin/Chettri. Similarly, non-Hindu women have higher odds of unmet need for FP, unmet need for limiting and spacing compared to their Hindu counterparts, which also findings aligns with a study conducted in Bihar, India that revealed differences based on ethnicity and religion^28^. Religious restrictions could potentially serve as a primary factor for the non-adoption of family planning methods.

Multiple previous studies have revealed that women with advanced education are less likely to have unmet need for spacing, limiting and family planning as evidenced across various global contexts^23,24,29,30^. However, our study reveals that women with basic and secondary education were more likely to have unmet need for FP compared to those who have no formal education. This could be because women with higher educational level have better understanding of the menstruation cycle and unsafe periods, as well as may have more concerns relating to potential side effects of modern contraceptives, which is corroborated by the findings that rhythm and withdrawal methods are more common among women having basic, secondary level or higher education compared to those who had no formal education.

Women belonging from wealthier households tend to have lower levels of unmet need for family planning, unmet need for spacing and limiting. This finding echoes findings from earlier studies conducted in diverse geographical regions, such as Pakistan^31^, Nigeria^32^, and Sub-Saharan Africa^33^. A plausible explanation for this pattern is the enhanced accessibility of modern contraceptive methods, as well as increased empowerment and autonomy among women from wealthier households as compared from women from poorer households.

In this study, parity emerged as a contributing factor of unmet need for family planning. Notably, women who had given birth to two or more children were more likely to have unmet need for family planning in contrast to those who had never given birth. These findings align harmoniously with results from other studies^23,34,35^. This phenomenon could potentially be attributed to the notion that, as the number of children a woman bears increases, the more likely the women will limit or space the number of children she will have.

Moreover, the study unveiled that woman who had no access to media exhibited a higher likelihood of unmet need for family planning in comparison to their counterparts who had access to media. These findings align with findings from prior studies conducted in Mozambique^36^ as well as Ethiopia^37–39^. Plausible justification for this lies in the potential of media access to counter prevailing misconceptions that impact contraceptive utilization through the facilitation of behavioral transformations.

The study shows a promising opportunity to enhance access to family planning services but calls for specific and targeted actions. A key approach could be to focus on strategy that generates demand, aiming to empower healthcare seeking behaviors, particularly among marginalized women. Policy makers and managers could design programs that provide women with personalized counseling on the full range of contraceptive options which allows them to choose methods that align with their unique situations and aspirations, with flexibility to switch methods as needed. These efforts could extend to both men and women, creating an environment where both partners feel comfortable seeking support and encouraging open conversations about family planning. By building upon the existing policies, Nepal has the potential to drive considerable progress in reducing unmet need for family planning, progressing towards achieving sustainable development goals.

## Strengths and Limitations

There are several strengths of this study. First, this study utilized a nationally representative dataset to examine the factors associated with unmet need for family planning among currently married women in Nepal. Thus, the findings of this study can be generalized throughout Nepal. Second, the results from this study used appropriate techniques and results perfectly agree with the past research findings. Third, we used weighted analysis to account complex survey design of the NDHS survey. However, there are some limitations. First this study is only limited to the demand side factors and does not consider the supply side factors like the availability of family planning methods or counseling, provider competency, or quality of care. Second, there are chances of recall bias in the study as women may give socially acceptable responses and may find it difficult to recall past experiences. Third, Cross-sectional nature of the data limits the causality between dependent and independent factors.

## Conclusion

The study found that many factors shape the unmet need for family planning among currently married women in Nepal. Findings uncovered intriguing patterns that cut across age groups, education levels, wealth differences and belonging to marginalized population. These findings underscore the importance of crafting solutions that fit the unique situations of these women. Furthermore, enhancing the grassroots-level women development initiatives becomes essential. This entails bolstering the efforts of local advocates who can identify obstacles to accessing services and facilitate the availability of healthcare through the dissemination of crucial health-related information to achieve the target set forth in the sustainable development goal.

## Acknowledgement

We would like to acknowledge “The DHS program” for providing dataset for further analysis and express our gratitude towards all who directly and indirectly supported and motivated us for this study.

## Data availability statement

The data are available publicly in the open-access repository. The data can be downloaded from the official website of ‘The Demographic and Health Surveys’ program. (https://dhsprogram.com/data/dataset/Nepal_Standard-DHS_2022.cfm?flag=0)

## Contribution

SPKC was responsible for conceptualization, data acquisition, formal analysis, methodology, validation, writing-original draft, and writing review & editing. BA was responsible for formal analysis, methodology, validation, writing-original draft and writing review and editing. ARP was responsible for methodology, validation, writing-original draft, and writing review & editing. MP, SK, SG, BL, SR, SS, BPD, DJ and GG were responsible for validation, writing-original the draft, and writing-review & editing. SCB were responsible for, supervision validation and writing review & editing. SPKC is the guarantor of the manuscript.

## References

1. Kassim M, Ndumbaro F. Factors affecting family planning literacy among women of childbearing age in the rural Lake zone, Tanzania. BMC Public Health 2022; 22: 1–11.

2. Prata N, Fraser A, Huchko MJ, et al. Women’s empowerment and family planning: A review of the literature. J Biosoc Sci 2017; 49: 713–743.

3. Dejene H, Abera M, Tadele A. Unmet need for family planning and associated factors among married women attending anti-retroviral treatment clinics in Dire Dawa City, Eastern Ethiopia. PLoS One 2021; 16: 1–15.

4. Haakenstad A, Angelino O, Irvine CMS, et al. Measuring contraceptive method mix, prevalence, and demand satisfied by age and marital status in 204 countries and territories, 1970–2019: a systematic analysis for the Global Burden of Disease Study 2019. Lancet 2022; 400: 295–327.

5. Adde KS, Dickson KS, Ameyaw EK, et al. Contraception needs and pregnancy termination in sub-Saharan Africa: a multilevel analysis of demographic and health survey data. Reprod Health 2021; 18: 1–9.

6. Wai MM, Bjertness E, Stigum H, et al. Unmet need for family planning among urban and rural married women in yangon region, myanmar—a cross-sectional study. Int J Environ Res Public Health; 16. Epub ahead of print 2019. DOI: 10.3390/ijerph16193742.

7. Casterline JB, Sinding SW. Unmet need for family planning in developing countries and implications for population policy. Popul Dev Rev 2000; 26: 691–723.

8. Mosuse MA, Gadeyne S. Prevalence and factors associated with unmet need for family planning among women of reproductive age (15–49) in the Democratic Republic of Congo: A multilevel mixed-effects analysis. PLoS One 2022; 17: 1–16.

9. Smith M, Keckley P. Adding it up: The costs and Benefits of investing in family planning and maternal and newborn health. New york Guttmacher Inst United Nations Popul Fund 2009; 91: 64–66.

10. Gahungu J, Vahdaninia M, Regmi PR. The unmet needs for modern family planning methods among postpartum women in Sub-Saharan Africa: a systematic review of the literature. Reprod Health 2021; 18: 1–15.

11. Mehata S, Paudel YR, Mehta R, et al. Unmet need for family planning in nepal during the first two years postpartum. Biomed Res Int; 2014. Epub ahead of print 2014. DOI: 10.1155/2014/649567.

12. Bhusal CK, Bhattarai S. Factors Affecting Unmet Need of Family Planning Among Married Tharu Women of Dang District, Nepal. Int J Reprod Med 2018; 2018: 9312687.

13. Tuladhar JM. Effect of family planning availability and accessibility on Contraceptive use in Nepal. Stud Fam Plann 1987; 18: 49–53.

14. Gubhaju B. Barriers to sustained use of contraception in Nepal: Quality of care, socioeconomic status, and method-related factors. Biodemography Soc Biol 2009; 55: 52–70.

15. Darroch JE, Singh S. Trends in contraceptive need and use in developing countries in 2003, 2008, and 2012: An analysis of national surveys. Lancet 2013; 381: 1756–1762.

16. Ministry of Health and Population [Nepal], New ERA I. Nepal Demographic and Health Survey 2022. Kathmandu, Nepal, 2023.

17. USAID. STAT Compiler: The DHS Programme, https://www.statcompiler.com/en/ (2023, accessed 16 August 2023).

18. Bhattathiry M, Ethirajan N. Unmet need for family planning among married women of reproductive age group in urban Tamil Nadu. J Fam Community Med 2014; 21: 53–57.

19. Genet E, Abeje G, Ejigu T. Determinants of unmet need for family planning among currently married women in Dangila town administration, Awi Zone, Amhara regional state; A cross sectional study. Reprod Health 2015; 12: 1–5.

20. Getaneh T, Negesse A, Dessie G, et al. Predictors of unmet need for family planning in Ethiopia 2019: a systematic review and meta analysis. Arch Public Heal 2020; 78: 1–11.

21. Anand B, Singh J, Mohi M. Study of Unmet Need for Family Planning In Immunisation Clinic of A Teaching Hospital at Patiala, India. Internet J Heal 2012; 11: 2–5.

22. Devi DR, Rastogi SR, Retherford RD. Unmet Need for Family Planning in Uttar Pradesh. Int Inst Popul Sci 1996; 1–27.

23. Hailemariam A, Haddis F. Factors Affecting Unmet Need for Family Planning In Southern Nations, Nationalities and Peoples Region, Ethiopia. Ethiop J Health Sci 2011; 21: 77–89.

24. Ojakaa D. Trends and Determinants of Unmet Need for Family Planning in Kenya. Macro Int.

25. Westoff BCF. The Potential Demand for Family Planning▫: A New Measure of Unmet Need and Estimates For Five Latin American Countries. Int Fam Plann Perspect 1988; 14: 45–53.

26. Woldemicael G, Beaujot R. Currently married women with an unmet need for contraception in Eritrea: Profile and determinants. Can Stud Popul 2011; 38: 61–81.

27. Teferi HM, Schröders J. Contributing factors for urban-rural inequalities in unmet need for family planning among reproductive-aged women in Ethiopia: a Blinder-Oaxaca decomposition analysis. BMC Womens Health 2023; 23: 1–10.

28. Pal A, Yadav J. S, et al. Factors associated with unmet need of family planning in Bihar, India: a spatial and multilevel analysis. Int J Reprod Contraception, Obstet Gynecol 2018; 7: 3638.

29. Acacio-Claro PJB, Borja MP. Addressing unmet need: Potential for increasing contraceptive prevalence in the Philippines. Asia-Pacific Popul J 2010; 25: 5–26.

30. Ali AAA, Okud A. The contribution of fulfilling the unmet need for family planning. Factors Affect unmet need Fam Plan East Sudan 2013; 13: 5.

31. Asif MF, Pervaiz Z. Socio-demographic determinants of unmet need for family planning among married women in Pakistan. BMC Public Health 2019; 19: 1–8.

32. Fagbamigbe AF, Afolabi RF, Idemudia ES. Demand and Unmet Needs of Contraception Among Sexually Active In-Union Women in Nigeria: Distribution, Associated Characteristics, Barriers, and Program Implications. SAGE Open; 8. Epub ahead of print 2018. DOI: 10.1177/2158244017754023.

33. Ahinkorah BO. Predictors of unmet need for contraception among adolescent girls and young women in selected high fertility countries in subsaharan africa: A multilevel mixed effects analysis. PLoS One 2020; 15: 1–15.

34. Gebre G, Birhan N, Gebreslasie K. Prevalence and factors associated with unmet need for family planning among the currently married reproductive age women in shire-Enda-Slassie, northern west of Tigray, Ethiopia 2015: A community based cross-sectional study. Pan Afr Med J 2016; 23: 1–9.

35. Moore Z, Pfitzer A, Gubin R, et al. Missed opportunities for family planning: An analysis of pregnancy risk and contraceptive method use among postpartum women in 21 low- and middle-income countries. Contraception 2015; 92: 31–39.

36. Yaya S, Idriss-Wheeler D, Uthman OA, et al. Determinants of unmet need for family planning in Gambia & Mozambique: implications for women’s health. BMC Womens Health 2021; 21: 1–8.

37. Dingeta T, Oljira L, Worku A, et al. <p>Unmet Need for Contraception Among Young Married Women in Eastern Ethiopia</p>. Open Access J Contracept 2019; Volume 10: 89–101.

38. Solomon T, Nigatu M, Gebrehiwot TT, et al. Unmet need for family planning and associated factors among currently married reproductive age women in Tiro Afeta District, South West Ethiopia, 2017: Cross-sectional study. BMC Womens Health 2019; 19: 1–9.

39. Alem AZ, Agegnehu CD. Magnitude and associated factors of unmet need for family planning among rural women in Ethiopia: A multilevel cross-sectional analysis. BMJ Open 2021; 11: 1–11.

